# Beyond the Spike: identification of viral targets of the antibody responses to SARS-CoV-2 in COVID-19 patients

**DOI:** 10.1101/2020.04.30.20085670

**Authors:** Asmaa Hachim, Niloufar Kavian, Carolyn A Cohen, Alex WH Chin, Daniel KW Chu, Chris KP Mok, Owen TY Tsang, Yiu Cheong Yeung, Ranawaka APM Perera, Leo LM Poon, Malik JS Peiris, Sophie A Valkenburg

## Abstract

**Background:** The SARS-CoV-2 virus emerged in December 2019 and caused a pandemic associated with a spectrum of COVID-19 disease ranging from asymptomatic to lethal infection. Serology testing is important for diagnosis of infection, determining infection attack rates and immunity in the population. It also informs vaccine development. Although several serology tests are in use, improving their specificity and sensitivity for early diagnosis on the one hand and for detecting past infection for population-based studies, are priorities.

**Methods:** We evaluated the anti-SARS-CoV-2 antibody profiles to 15 SARS-CoV-2 antigens by cloning and expressing 15 open reading frames (ORFs) in mammalian cells and screened antibody responses to them in COVID-19 patients using the Luciferase Immunoprecipitation System (LIPS).

**Results:** The LIPS technique allowed us to detect antibody responses in COVID-19 patients to 11 of the 15 SARS-CoV-2 antigens tested, identifying novel immunogenic targets. This technique shows that antigens ORF3b and ORF8 allow detection of antibody early in infection in a specific manner and reveals the immuno-dominance of the N antigen in COVID-19 patients.

**Conclusion:** Our report provides an unbiased characterization of antibody responses to a range of SARS-CoV-2 antigens. The combination of 3 SARS-CoV-2 antibody LIPS assays, i.e. N, ORF3b, and ORF8, is sufficient to identify all COVID-19 patients of our cohort even at early time-points of illness, whilst Spike alone fails to do so. Furthermore, our study highlights the importance of investigating new immunogens NSP1, ORF3b, ORF7a and ORF8 which may mediate immune functions other than neutralization which may be beneficial or harmful to the patient.

## Introduction

The acute pandemic respiratory disease COVID-19 is caused by a novel coronavirus that belongs to the species *Severe acute respiratory syndrome-related coronavirus* (SARS-CoV-2) (1, 2). There are at least 70 vaccine candidates against COVID-19 in various stages of testing, development and human clinical trials (5). Urgency in vaccine development and deployment to mitigate the pandemic has left some fundamental immunological research questions outstanding, especially regarding the range of virus immunogens that elicit antibody responses, their kinetics, specificity, breadth, longevity and impact for long-term protection or immune-pathology.

Endemic human coronaviruses (HCoV) OC43, 229E, HKU1 and NL63 usually cause a mild ‘common cold’- like upper respiratory disease. SARS-CoV-2 belongs to the β-coronavirus genus which includes SARS-CoV which emerged in 2002, and Middle Eastern Respiratory Syndrome Coronavirus (MERS-CoV) which emerged in 2012, which are severe human diseases of zoonotic origin (4). As of 20th of April 2020, the World Health Organization has reported a total of 2,241,778 cases of COVID-19 worldwide and 152,551 deaths (5). Ongoing research has reported the full genome sequence (1), transcriptome (6) and the immune response to infection (7).

SARS-CoV-2 has 12 putative functional open reading frames (ORF) and shares 82% nucleotide homology with SARS-CoV (8). There are at least four structural proteins in SARS-CoV-2: Spike (S), Envelope (E), Membrane (M), Nucleocapsid (N). The trimer S protein is cleaved into S1, containing the receptor binding domain, and S2 subunits (8, 9), and S2 is further cleaved into S2’ to form the viral fusion peptide (6). The S protein is critical for viral entry and is a neutralising target, as it is with SARS-CoV and MERS-CoV and is a key target for diagnostic tests and vaccine development (10). The S1 subunit of SARS-CoV-2 shares about 70% identity with the SARS-CoV, whereas the identity of the S2 subunit is up to 99% with some evidence of cross-reactivity between the viruses (11). Apart from these structural proteins, the SARS-CoV-2 genome encodes for around 20 putative non-structural proteins (8). ORF1a/b encodes for a large polyprotein that is proteolytically cleaved into 16 non-structural proteins (NSP1–16). Extra ORFs, such as ORF3a, 3b, 6, 7a, 7b, 8 and 10 may encode for proteins but their functions are unknown.

Encouragingly, reports have shown that SARS-CoV-2 patients develop neutralising and high titer S1-specific antibody responses (11), and robust T and B cellular responses (12). However, the magnitude of the antibody responses appears related to the clinical severity of COVID-19 disease. In one recent report 10 of 175 confirmed patients did not develop a neutralising SARS-CoV-2 specific antibody response (by pseudo particle assay), but binding antibodies were still detected (by ELISA) (13). In patients with MERS (14) it has been shown that viral load and severe disease correlates with the magnitude of the antibody response, however mild diseases (in 4 of 6 mild subjects) may not lead to detectable antibody responses. Furthermore, MERS-CoV seropositive dromedary camels can still be re-infected in the presence of high antibody titers (15). Similarly, there are reports of waning of S1-specific and neutralising antibodies in SARS-CoV infection (16). These instances of low or no antibody responses by traditional serological approaches may lead to an underestimation of asymptomatic and mild infection, and threaten the success of a potential vaccine that targets the S1 alone. Therefore, a broader landscape of antibody responses to a range of viral proteins needs to be assessed to better detect the immunogenicity of SARS-CoV-2 infection to improve the understanding of pathogenesis and immunity.

The Luciferase Immunoprecipitation system (LIPS) assay allows for a comprehensive understanding of the antibody immune response against infectious agents (17) by the expression of any protein or antigen as a recombinant Renilla luciferase (Ruc)-antigen fusion. The LIPS technique has previously been used to distinguish infected versus vaccinated cases for influenza due to the presence of antibodies against non-structural proteins (18), and to characterize human infections by zoonotic spill over from bat viruses (19). In this study, we used LIPS to assess the acute and convalescent antibody responses to a panel of 15 potential SARS-CoV-2 antigens: the 4 structural proteins (S, N, M and E), 3 S subunits (S1, S2, S2’), the 7 available ORFs (ORF3a, 3b, 6, 7a, 7b, 8 and 10) and 1 relevant NSP within ORF1a/b (NSP1) (20). Our data reports the most extensive landscape of antibody responses of COVID-19 patients reported to date.

## RESULTS

We made a panel of fifteen SARS-CoV-2 ORFs as *Renilla* luciferase-antigen fusion proteins to assess the humoral immune responses in 15 COVID-19 infected patients from Hong Kong compared with a panel of healthy negative controls using LIPS. The four SARS-CoV-2 structural proteins show high amino-acid sequence homology with SARS-CoV but not with other human coronaviruses responsible for common colds (Supplementary Table 1). We detected significantly higher antibody responses to the full Spike protein (S), the Nucleoprotein (N) and the Membrane protein (M) in COVID-19 patients compared to healthy negative controls (p <0.0001, p=0.023, and p=0.0116 respectively, Figure 1bcd). COVID-19 patients did not show any increased production of Envelope (E) antibodies (Figure 1e) compared to healthy negative controls (p=0.0591, Figure 1d).

**Figure 1.**
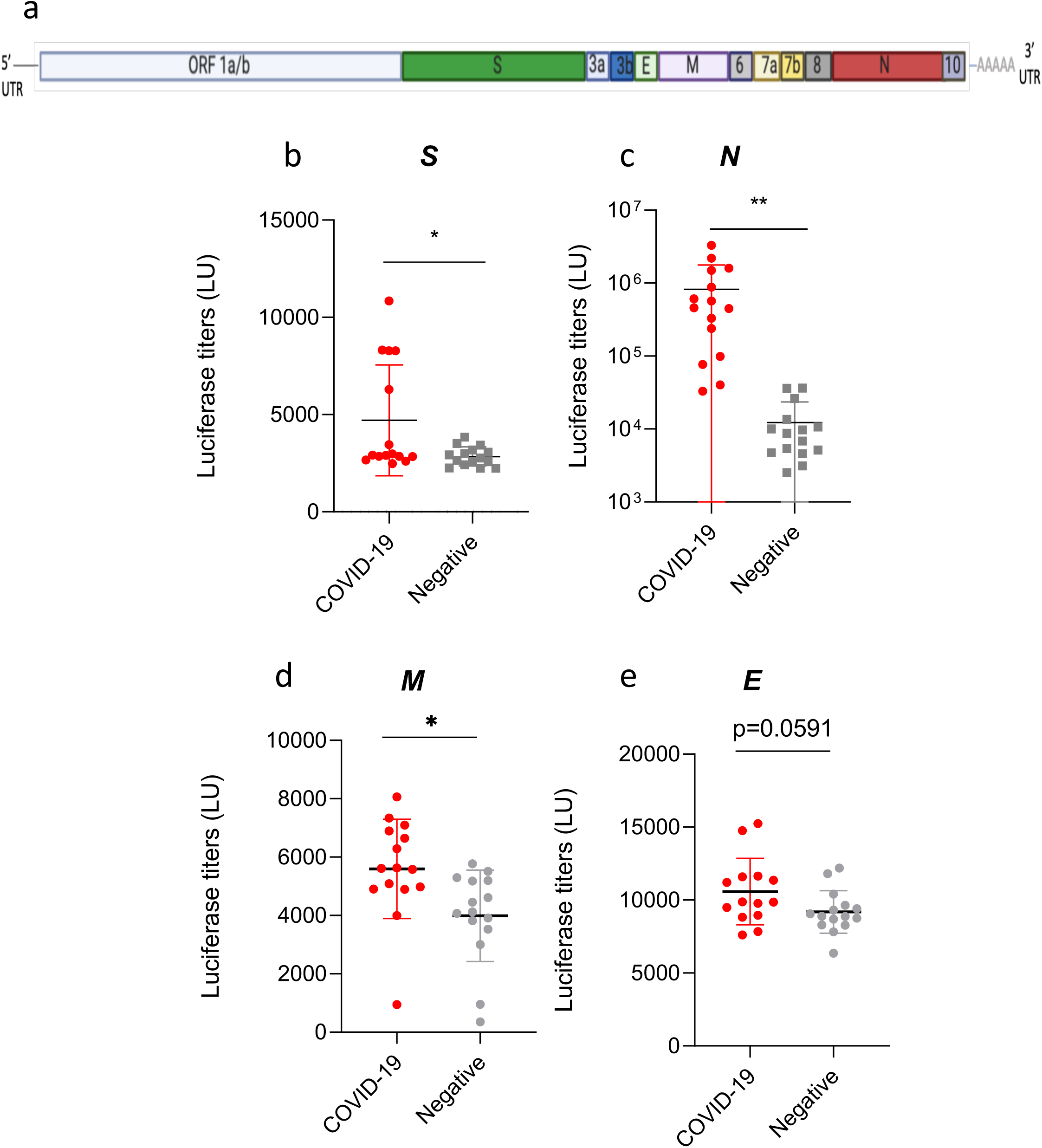
Detection of SARS-CoV-2 structural proteins antibodies by LIPS. (a) SARS-CoV-2 genome ORF organisation *(not to scale)*. (b-e) Antibodies against the four SARS-CoV-2 structural proteins Spike (S), Nucleocapsid (N), Membrane (M), Envelope (E) were measured by LIPS from COVID-19 patients, and age matched negative controls. Data represents the mean +/− stdev, and individual responses (n=15). Background values were subtracted. Experiments were repeated twice. P values were calculated using the Mann-Whitney U test. * shows statistical significance between COVID-19 patients versus negative controls. ns, p=0.0591 *p<0.05, **p<0.01.

We confirmed our results by traditional IgG ELISA, which correlated well with LIPS LU responses (R2=0.5289) for full-S (Supplementary figure 1ab). N-specific IgG by ELISA is also elevated in COVID-19 patients (Supplementary figure 1c) but does not correlate strongly with LIPS results (Supplementary figure 1d).

The S1 subunit, a key virus immunogen, detected significantly higher antibodies in COVID-19 patients than negative controls (5191+/−1469 LU versus 4003+/−1062 LU, p=0.0288, Figure 2a). Interestingly, there was no difference in the antibodies to S2 in the LIPS assay between COVID-19 patients and negative controls (p=0.5683). Antibodies to the S2’ cleaved subunit were significantly higher in COVID-19 patients (p=0.0391 for S2’, Figure 2) but the overall in antibody levels between the groups was considerable. Patients with higher SARS-CoV-2 micro neutralization (MN) titers (>160 reciprocal serum dilution) also had higher responses towards full-S by LIPS, whilst there was no difference observed in LIPS responses to subunits S1, S2 and S2’ in high or low MN COVID-19 patients (Figure 2c, p=0.0049).

**Figure 2.**
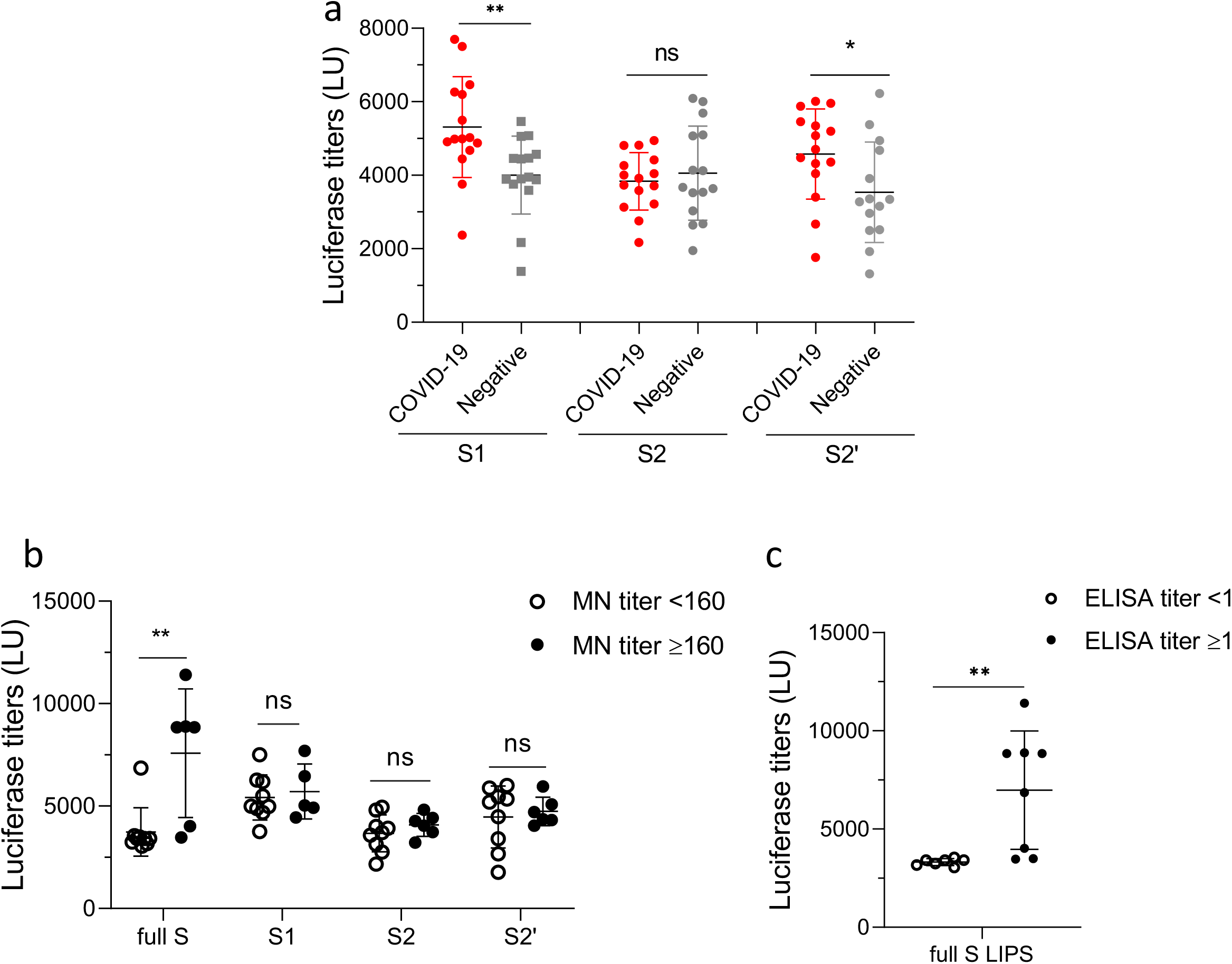
LIPS detection of antibody levels to the SARS-CoV-2 S protein subunits. (a) Antibodies against the S subunits S1, S2, and S2’ by LIPS from COVID-19 patients and negative controls. (b) Full S, S1, S2, S2’ antibodies LIPS responses in COVID-19 patients with low microneutralization (MN) titers (<160) versus high MN titers (≥160). (c) Full S antibodies LIPS responses in COVID-19 patients with low ELISA S IgG responses (<1) versus high ELISA IgG responses (≥1). Data represents the mean +/− stdev, and individual responses (n=15). Background values were subtracted. Experiments were repeated twice. P values were calculated using the Mann-Whitney U test. * shows statistical significance between COVID-19 patients versus negative controls. *p<0.05, **p<0.01.

We next investigated the presence of antibodies specific to previously uncharacterized ORFS to the SARS-CoV-2. As we could not produce the full ORF1ab due to its extended length (>21,000 bp, (1)), we cloned and expressed a representative antigen, non-structural protein 1 (NSP1). Bioinformatic predictions have revealed a putative role for NSP1 in suppressing the antiviral host response (8).

We used LIPS to detect antibodies specific to NSP1, ORF3a, ORF3b, ORF6, ORF7a, ORF7b, ORF8 and ORF10 (Figure 3). COVID-19 patients had significantly higher NSP1-antibody levels compared to negative controls (mean of 5301+/−854.7 LU versus 3683+/−726.4 LU, p<0.0001, Figure 3a). Furthermore, significantly higher antibody levels in COVID-19 patients were detected towards ORF3a, ORF3b, ORF7a, ORF7b and ORF8 (p=0.0302, p<0.0001, p<0.0001, p=0.0019, p<0.0001, Figure 3b, c, e, f, g). The largest difference between COVID-19 and negative controls in mean antibody signals among the ORFs was observed for ORF3b (7712+/−2947 LU versus 3599+/−1029 LU) and ORF8 (16933+/−7489 LU versus 5440+/−1096 LU) (Figure 3c and g). Meanwhile there was no significant difference between COVID-19 patients and negative controls for ORF6 and ORF10 antigens (Figure 3d and h).

Globally, among the antibody responses tested in our LIPS assay, we detected significant levels of antibodies specific to 11 antigens: N, M, S, S1, S2’, NSP1, ORF3a, ORF3b, ORF7a, ORF7b and ORF6 in the COVID-19 population (Table 1) at all time-points of infection from day 4 to day 22 (see below in Figure 4 for early time-point detection). We did not detect a significant production of antibodies to 4 of the SARS-CoV-2 antigens tested: E, S2, ORF6 and ORF10 proteins.

**Figure 3.**
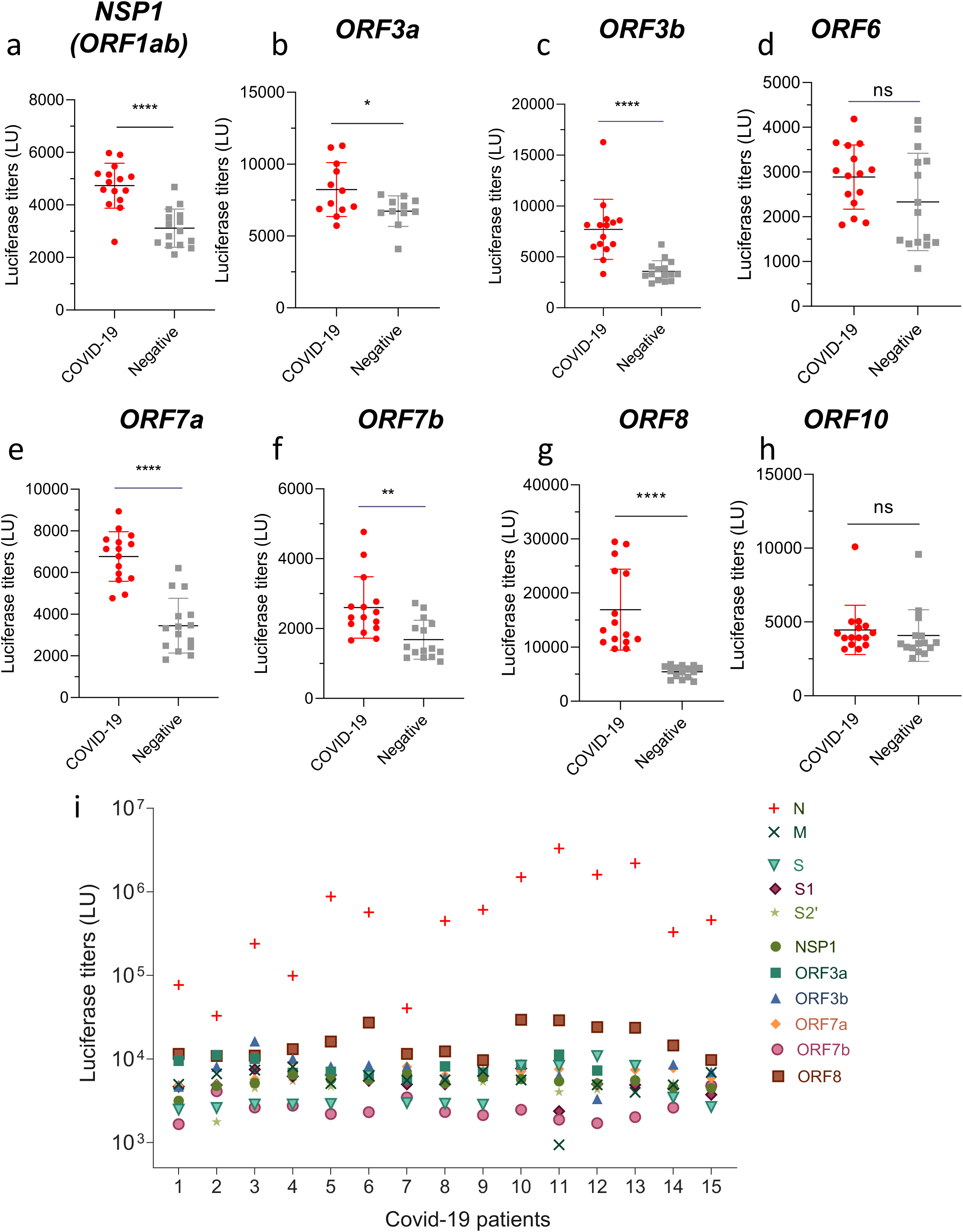
Detection of SARS-CoV-2 non-structural proteins antibodies by LIPS. (a-h) Antibodies against NSP1 (in ORF1ab), and other ORFs (ORF3a, 3b, 6, 7a, 7b, 8 and 10) were measured by LIPS to cover all the ORFs of the virus. Data represents the mean +/− stdev, and individual responses (n=15). (i) Global individual immune responses detected by LIPS in the 15 COVID-19 patients for the 11 relevant antigens. Background values were subtracted. Experiments were repeated twice. P values were calculated using the Mann-Whitney U test. * shows statistical significance between COVID-19 patients versus negative controls. *p<0.05, **p<0.01, ***p<0.001, ****p<0.0001.

**Figure 4.**
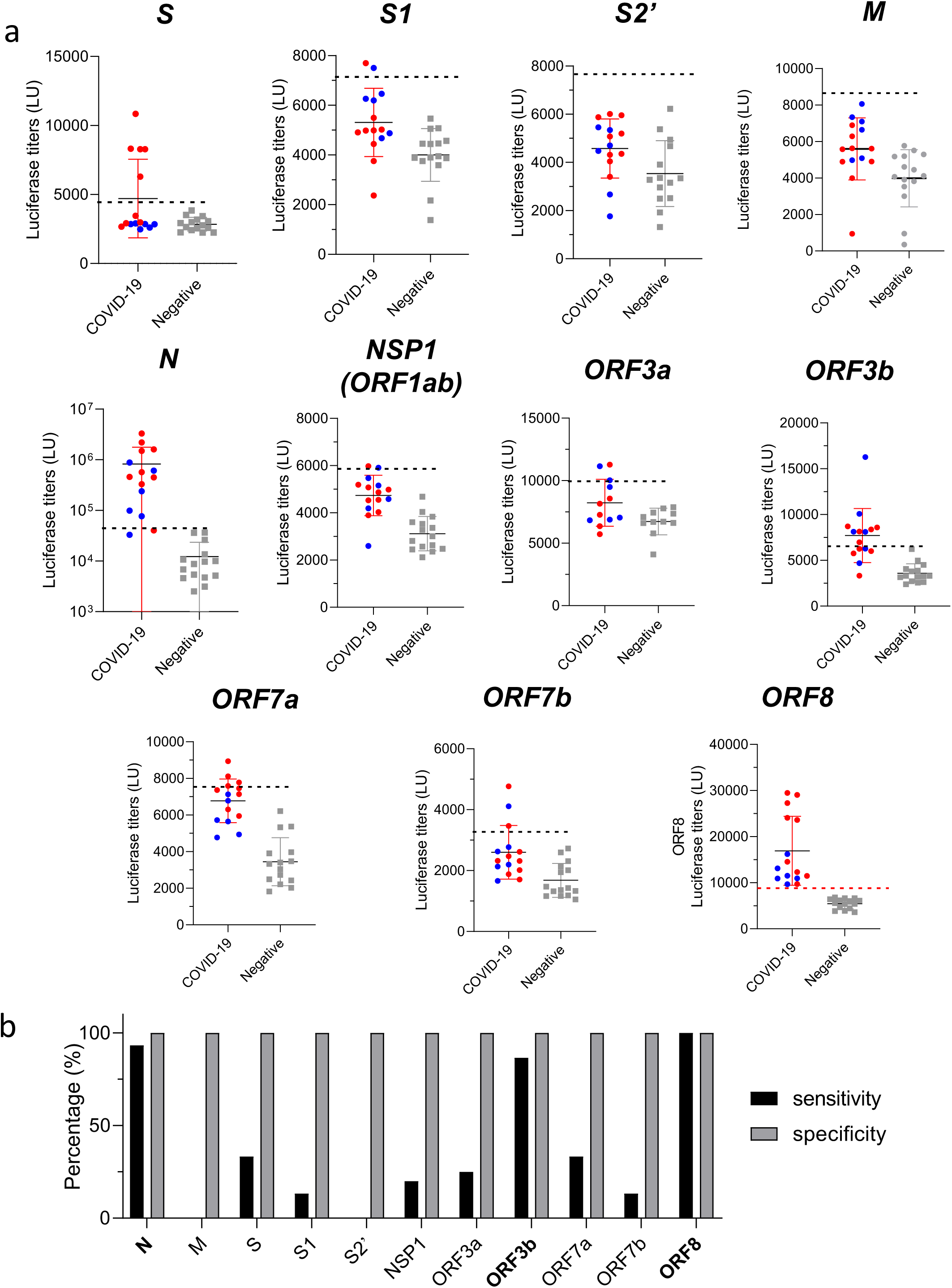
Sensitivity of LIPS tests for potential diagnostic use. (a) Cut-off values for the 11 relevant LIPS tests (dotted line) were calculated as mean of negatives + 3 Stdev. Early time-points (<day 14) COVID-19 samples are shown by the blue dots. Data represents the mean +/− stdev, and individual responses (n=15). The red dotted line in ORF8 represents a 100% sensitivity and specificity. Background values were subtracted. Experiments were repeated twice. (b) Sensitivity and specificity performances of the 11 LIPS tests.

**Table 1.** List of ORFs produced as Renilla-Antigen fusion protein

| Antigen | Full name | Patient response |
| --- | --- | --- |
| N | Nucleocapsid | Yes |
| M | Membrane | Yes |
| E | Envelope | No |
| S | Spike | Yes |
| S1 | Spike S1 subunit | Yes |
| S2 | Spike S2 subunit | No |
| S2' | Spike S2 subunit | Yes |
| NSP1 (in ORF1a/b) | - | Yes |
| ORF3a | - | Yes |
| ORF3b | - | Yes |
| ORF6 | - | No |
| ORF7a | - | Yes |
| ORF7b | - | Yes |
| ORF8 | - | Yes |
| ORF10 | - | No |

**Table 2.**
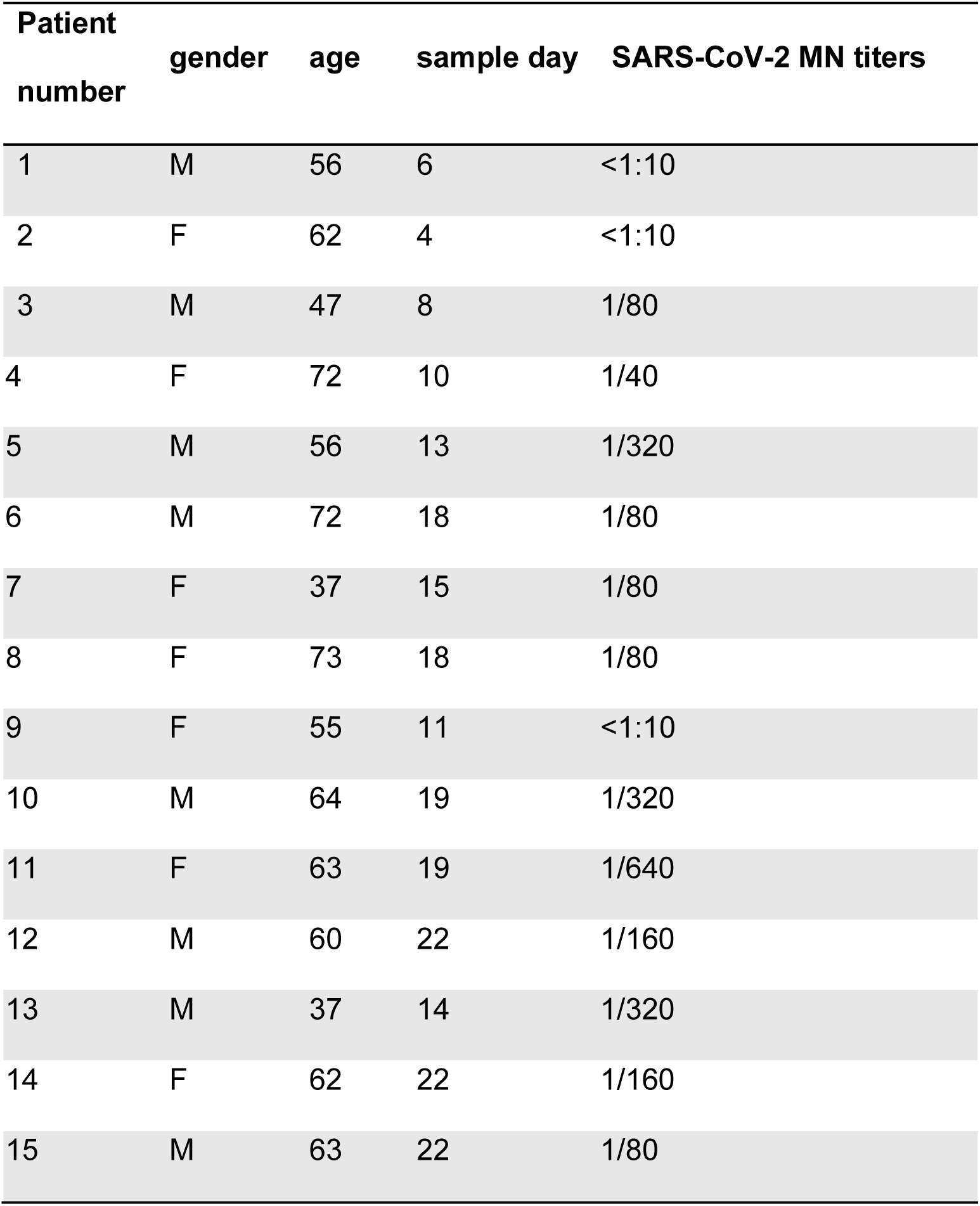
Hong Kong COVID-583 19 infected patient information

| Patient<br>number | gender | age | sample day | SARS-CoV-2 MN titers |
| --- | --- | --- | --- | --- |
| 1 | M | 56 | 6 | <1:10 |
| 2 | F | 62 | 4 | <1:10 |
| 3 | M | 47 | 8 | 1/80 |
| 4 | F | 72 | 10 | 1/40 |
| 5 | M | 56 | 13 | 1/320 |
| 6 | M | 72 | 18 | 1/80 |
| 7 | F | 37 | 15 | 1/80 |
| 8 | F | 73 | 18 | 1/80 |
| 9 | F | 55 | 11 | <1:10 |
| 10 | M | 64 | 19 | 1/320 |
| 11 | F | 63 | 19 | 1/640 |
| 12 | M | 60 | 22 | 1/160 |
| 13 | M | 37 | 14 | 1/320 |
| 14 | F | 62 | 22 | 1/160 |
| 15 | M | 63 | 22 | 1/80 |

Comparison of the global SARS-CoV-2 antibody responses from 15 COVID-19 patients reveals that anti-N antibodies dominate the humoral response detected by LIPS (Figure 3i), whilst other antigens make lower and similar contributions to the magnitude anti-SARS-Cov-2 antibody responses (Figure 5a).

**Figure 5.**
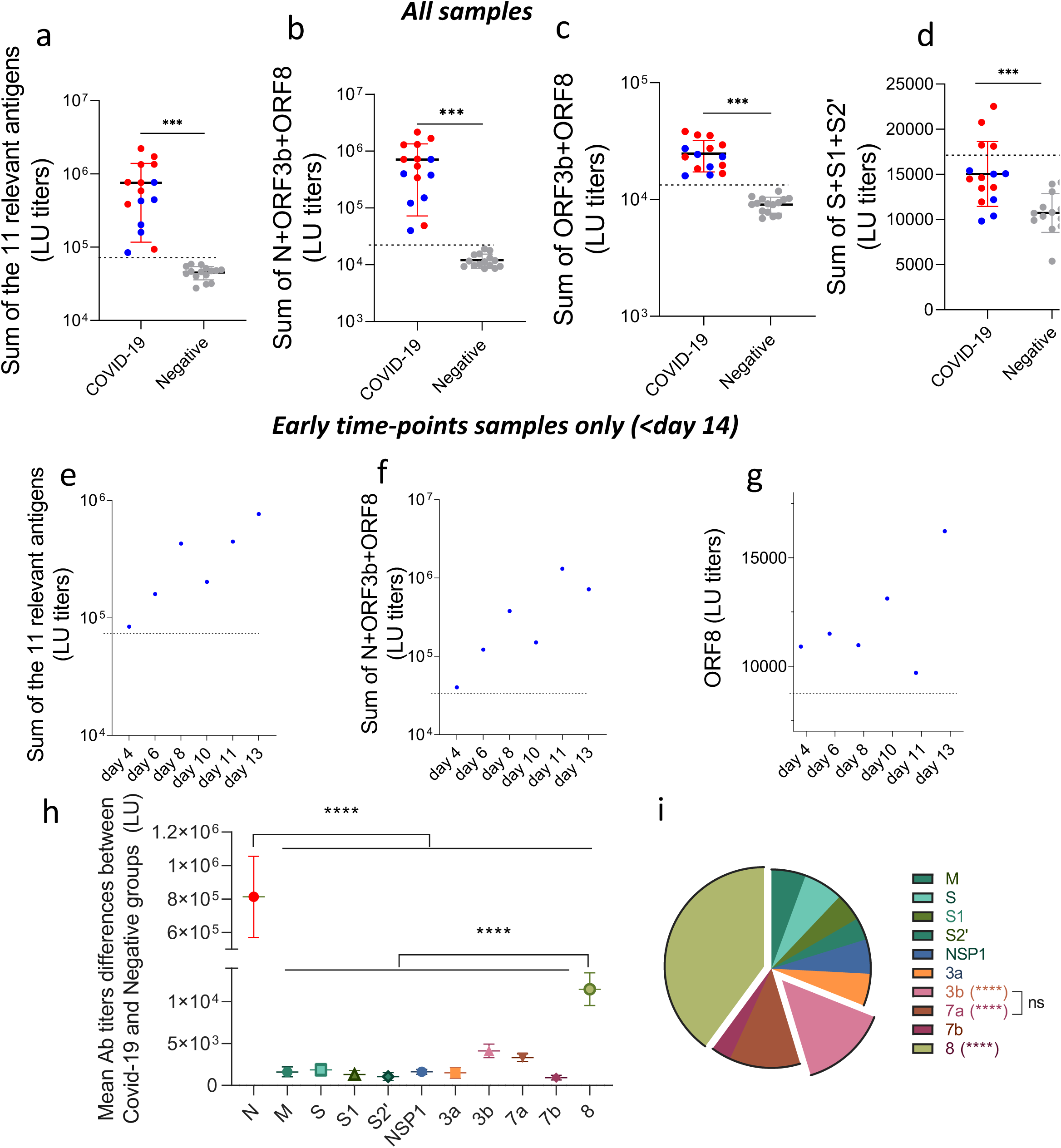
Combining LIPS tests as a diagnostic tool for COVID-19. Cumulative antibody LIPS levels to the SARS-CoV-2 antigens in COVID-19 patients and negative controls, for (a) 11 relevant antigens (N, M, S, S1, S2’, NSP1, ORF3a, ORF3b, ORF7a, ORF7b, ORF10) (b) three most sensitive antigens (N, ORF3b, and ORF8), (c) selective antigens (ORF3b, and ORF8), and (d) 3 S relevant antigens (S, S1, S2’). (e-f) Detection of early time-points patients only with the sum of (e) 11 relevant Antigens and (f) three most sensitive antigens (N+ORF3b+ORF8). Data represents the mean +/− stdev, and individual responses (n=15). The cut-off value of the sums is shown by the dotted line and was based on the mean + three stdev of the negative control group. Blue dots represents COVID-19 patients data points prior to day 14. (h-i) Differences of the antibody response means between COVID-19 and negative control populations as (h) scatter plot including the 11 relevant antigens, and as (i) pie chart excluding N. Each symbol represents the mean +/− stdev of the differences between COVID-19 and negative groups, for the 11 relevant antibody levels in light units (LU). *** p<0.001, **** p<0.0001 versus all other antigens, otherwise indicated.

### Absence of sex- and age- effect in our cohort

Epidemiological data from many countries suggest that men develop more severe COVID-19 related symptoms than women (21). We therefore evaluated the effect of gender in our cohort of COVID-19 patients for antibody responses and found no difference across all antigenic targets tested (p=0.6289, Supplementary Figure 2a). Similarly, the SARS-CoV-2 antibody response was comparable between patients aged below and above 60 years old (p=0.9363, Supplementary Figure 2b).

### Combined antigen test panels as a potential diagnostic tool for COVID-19

We found significantly higher levels of 11 of the 15 antigens tested in the COVID-19 populations. Next, their ability to correctly identify COVID-19 patients was determined. Therefore, a cut-off value of LIPS antibody signals, based on the mean plus three standard deviations of the healthy negative control group (22–24), was calculated for each of these 11 antigens, along with the sensitivity and specificity of each test (Table 3, and Figure 4). The cut-off value of all 11 tests showed a high specificity of the SARS-CoV-2 LIPS assays with no samples from the negative control group being above the cut-off. On the other hand, 8 antigens showed a low sensitivity (below 75% for M, S, S1, S2’, NSP1, ORF3a, ORF7a, ORF7b, Figure 4b), hence being insufficient to correctly identify all the COVID-19 patients with high rates of false negatives (Table 3), limiting their use for LIPS diagnostics. On the other hand, the N, ORF3b and ORF8 antigens showed good sensitivity levels of 93.3%, 86.6% and 100% respectively (Table 3). Of note, ORF8 is the only of the 11 tests to correctly identify all COVID-19 patients including at day 4 after onset of illness (Figure 4a).

**Table 3.** Cut-off value, sensitivity, specificity, and Positive/Negative Predictive Values for: each of the 11 relevant SARS-CoV-2 antigens tests, as well as the sums of the 11 relevant antigens, N+ORF3b+ORF8, ORF3b+ORF8, and S+S1+S2’ tests. o

| Antigen | Cut-off<br>value | sensitivity | specificity | Positive predictive<br>value (PPV) | Negative predictive<br>value (NPV) |
| --- | --- | --- | --- | --- | --- |
| N | 46133 | 93.3% | 100% | 100% | 93.75% |
| M | 8684 | 0% | 100% | 0% | 50% |
| S | 4333 | 33.3% | 100% | 100% | 60% |
| S1 | 7189 | 13.3% | 100% | 100% | 53.57% |
| S2' | 7635 | 0% | 100% | 0% | 50% |
| NSP1 | 5862 | 20% | 100% | 100% | 55.56% |
| ORF3a | 9925 | 25% | 100% | 100% | 55.56% |
| ORF3b | 6686 | 86.6% | 100% | 100% | 88.24% |
| ORF7a | 7394 | 33.3% | 100% | 100% | 60% |
| ORF7b | 3355 | 13.3% | 100% | 100% | 53.57% |
| ORF8 | 8728 | 100% | 100% | 100% | 100% |
| Sum of the 11<br>relevant tests | 73323 | 100% | 100% | 100% | 100% |
| Sum of<br>N+ORF3b+ORF8 | 22101 | 100% | 100% | 100% | 100% |
| Sum of<br>ORF3b+ORF8 | 13280 | 100% | 100% | 100% | 100% |
| Sum of S+S1+S2' | 17134 | 26.67% | 100% | 100% | 57.69% |

As many LIPS tests showed low sensitivity, we then used an approach based on the combination of LIPS antibody LU signals for the 11 separate SARS-CoV-2 antigen tests to efficiently detect SARS-CoV-2 exposure in this population (22–24). This approach of combining the LU values from the 11 LIPS tests increased the sensitivity to 100% for the diagnosis of COVID-19 infections during acute infection (Figure 5a, blue dots for <14 days). Furthermore, combination of only 3 LIPS tests, those for anti-N, ORF3b and ORF8 antibodies, also has a 100% sensitivity and specificity (Figure 5b and Table 3). As ORF3b and ORF8 show the lowest homology to previous SARS-CoV among all the viral proteins (8), we also looked separately at the combination of responses towards ORF3b and ORF8 (Figure 5c). We observed the same trend as above with all COVID-19 patients having a combined score above the cut-off and all negative controls having a combined score below, and all early time-points being correctly detected (Figure 4c). By combining only the 3 relevant Spike protein LIPS tests (S, S1, S2’), the sensitivity of the LIPS test drastically decreased to 26.6% as only 4 of the 15 COVID-19 patients had a total combined LU above the cutoff (Figure 5d). Interestingly all samples from early time-points (day 4 to day 13) had LU signals under the cut-off value for S+S1+S2’ (blue dots, Figure 5d). The magnitude of the 11 relevant antigens, N+ORF3b+ORF8, as well as ORF3b+ORF8 responses significantly increase the detection of patients with early time-points samples (Figure 5e, f). Therefore, the combinational use of ORF3b and ORF8 tests alone could be sufficient to detect COVID-19 exposed subjects at any time-point of infection.

Because endemic HCoVs are ubiquitous, the negative control plasma samples are likely to have antibodies to a range of HCoVs. Sequence homology with other HCoV could result in the detection of cross-reactive antibodies by LIPS and reduce specificity of serological assays. Structural proteins of SARS-CoV-2 with other HCoV structural proteins only share 18 to 40% homology (Supplementary table 1), making potential cross-reactivity of existing antibodies unlikely and undetermined. Whilst SARS-Cov-2 and SARS-CoV, N and S share 94% and 76% conservation (Supplementary table 1), previous reports showed the lowest homology for ORF3b and ORF8 at 32% and 40% amino acid homology respectively (8), making them the most unique genes to SARS-CoV2.

To investigate specificity of the antibody responses to the panel of antigens, we subtracted the mean LU levels of healthy negative controls from the mean LU of COVID-19 infected patients and compared this difference across each antigen (Figure 5b). We found that this difference was significantly higher for the N-specific antibody response compared to all the other relevant antibody responses (M, S, S1, S2’, NSP1, ORF3a, ORF3b, ORF7a, ORF7b, and ORF8) (p<0.0001, Figure 5b), highlighting a possible dominance and specificity of N across the SARS-CoV-2 humoral immune responses. Besides N, the ORF8 and ORF3b also appear to be important antigenic targets (Figure 5h, i). Statistical analysis (ANOVA) revealed that ORF8 was significantly increased compared to all other antigens (p<0.0001 versus 10 remaining antigens: M, S, S1, S2’, NSP1, ORF3a, 3b, 7a, and 7b, Figure 5bc). Whilst results for ORF3b were significant against the remaining 8 antigens (M, S, S1, S2’, NSP1, ORF3a, 3b, and 7b), excluding ORF7a (Figure 5c). Therefore, ORF3b and ORF8 are newly identified as specific and unique antigenic targets.

## DISCUSSION

SARS-CoV-2 antibody testing is an important component of the options for diagnosis of recent and past COVID-19 infection. Antibody tests are crucial for determining infection attack rates in the population, population immunity and inform vaccine development. We report, for the first time, the detection of antibody responses directed against an extensive spectrum of the SARS-CoV-2 antigens. Several approaches have been developed to measure SARS-CoV-2-specific antibodies, including micro neutralization assays (virus or pseudo virus-based (25)), ELISA assays (11, 26), immunofluorescence (7), colloidal gold-based immunochromatographic assays (27), and peptide/protein microarray (28) (29). Using LIPS technology with crude lysates from transfected cells, we have screened all the structural proteins along with all the ORFs of the SARS-CoV-2 virus (NSP1 only for ORF1ab) to identify new and unique antigenic targets of the humoral immune response of COVID-19 patients.

Among the 15 proteins tested for antibody specificity, 11 antigens showed significantly higher responses in the COVID-19 patients compared to healthy prepandemic negative controls. For the Spike subunits, only antibodies to S1 and S2’ were elevated in COVID-19 patients by our LIPS test, and S2 responses were not significantly different from the control group. The trimer S conformation and maturation of viral particles by the cleavage of S2 during virus endocytosis to form the S2’ fusion peptide may result in a difference in their antigenicity. Patients with higher MN titer had correspondingly higher levels of full-Spike LIPS and ELISA results, establishing consistency among assays for the full-S protein. N antibody responses detected were also elevated in patients, but the correlation between LIPS and ELISA N antibody assays was lower.

We next cloned all the available ORFs of the virus ORF1ab (as NSP1 only), ORF3a, ORF3b, ORF6, ORF7a, ORF7b, ORF8 and ORF10, to acquire more extensive information of the immunogenic targets of the virus. We found that 6 of these 8 ORFs induced a humoral immune response in the patients (NSP1 (ORF1ab), ORF3a, ORF3b, ORF7a, ORF7b and ORF8). Furthermore, by calculating a cut-off value based on the mean of the negative population + 3 standard deviations, we could ascertain that only 3 out of these 11 antigens were useful in diagnostic tests with high performance: N, ORF3b, and ORF8. By using the combined results of several antibody signals and calculating a cut-off value for these responses, we found that the sum of the 11 relevant antibody tests was highly sensitive and specific. Moreover, combining only the 3 of the most informative and sensitive antigens, N, ORF3b and ORF8, we achieved a sensitivity and specificity of 100%, correctly identifying all the COVID-19 patients versus negative controls. A larger data set from COVID-19 patients and negative controls is needed to further refine and confirm the performance of our test (30).

The Spike protein is responsible for virus entry into host cells and is the main antigen that elicits neutralizing antibodies (10). Whilst we detected significant differences in the magnitude of responses by LIPS between patients and controls for S, S1 and S2’, these antigens did not show high sensitivity levels, especially for sera collected early post disease onset. The Spike protein is a trimer on the surface of virions, and the conformation of our Ruc-S antigen should be assessed by conformation dependent monoclonal antibodies to confirm whether the conformation of S protein in our assay is well retained for antibody binding and affinity (31). Further LIPS tests using the S RBD antigen alone are underway to assess the proportion of RBD-specific antibodies as an important target of neutralizing antibodies (31). The S protein also elicits non-neutralizing antibodies targeted to conserved epitopes (11), and among our cohort an absence of *in vitro* neutralization has been observed for some patients, especially at early time-points.

The combination of multiple antigens by LIPS beyond the Spike could be the basis for supplementary serological tests useful to determine SARS-CoV-2 exposure to overcome false negative results. We found that the single test detection of N, S or ORF3b antibodies at early time-points (day 4 to day 14) results in a high proportion of false negative results, whilst the minimal combination of N+ORF3b+ORF8 LIPS tests is highly sensitive and specific.

Importantly, ORF3b and ORF8 are the least identical proteins to SARS-CoV (8), and they do not exist in other strains of human coronaviruses. However very little is known about their function and expression. Previous reports found the ORF3b of SARS-CoV plays an important role in the interaction with the innate immune system through inhibition of type 1 Interferon synthesis (33). In SARS-CoV, ORF8 has been shown to accumulate in the Endoplasmic Reticulum and mediate cell death by autophagy (32). In SARS-CoV-2, the functions of these ORFs have yet to be determined. Importantly, ORF8 can only be found in human and bat SARS-like CoV (35), and in our results we observed very low background detection in negative control plasma resulting in highly specific results. However, there are recent reports of the deletion of ORF8 in a few Singaporean COVID-19 patients (34) but this is not reported elsewhere. Whilst ORF8-deletion SARS-CoV-2 viruses had reduced replicative fitness, this issue may undermine the utility of ORF8 alone in serological testing. In addition, NSP1 and ORF7a LIPS tests showed high significance for COVID-19 patients. Though little is known about their function, bioinformatic predictions reveal that both NSP1 and ORF7a could be involved in suppressing the antiviral host response (6, 8). Therefore, the combined use of multiple antigens that are sensitive and specific is needed for diagnostic serology.

Most commercially available or published serological tests use only the S antigen, with a few using both the S and N antigens (11, 36, 37). Using extensive testing of the virus antigens, we have shown that additional targets are important for early detection of antibody responses and identification of COVID-19 patients. Potential cross-reactivity of SARS-CoV-2 antibodies with other coronaviruses antibodies cannot be excluded, but several recent data report that this cross-reactivity is minimal (26). Therefore, the approach of combining several relevant antigens that are unique to SARS-CoV-2, and immunogenic boosting specific antibodies would overcome issues of cross-reactivity and increase the sensitivity of serological assays. Screening of additional negative pre-pandemic samples with our LIPS approach is needed to refine issues of cross-reactivity with other HCoV. The high orders of magnitude and range of antibody quantity measured by our SARS-CoV-2 LIPS assay is an advantage compared to ELISA Optical Density measurements. These advantages may help with the rapid screening of recovered COVID-19 patients for elevated antibodies to donate serum for the treatment of other patients by passive transfer (38).

Our study does not report on antibody functions and data on Fc mediated functions of SARS-CoV-2 antibodies are also still lacking (39). Therefore, Fc mediated functions of antibodies directed against internal proteins from ORF3b and ORF8 will be the focus of future studies. Furthermore, only IgG is bound to the protein A/G used in the LIPS assays, therefore the use of protein L that binds the light chain of all immunoglobulins could be of interest to detect IgM early responses in future investigations. The E, S2, ORF6 and ORF10 antibodies did not show elevated levels in COVID-19 patients in our assay, consistent with the findings of Wang H. *et al*. by microarray (29). Further confirmation is needed using additional experimental approaches before excluding their utility.

Our study is limited by small sample size and by the small number of sera collected in the first few days after onset of symptoms. The absence of paired samples from multiple time-points for the same patients is also a limitation. Longitudinal studies are needed to further assess kinetics of antibody responses, especially to identify early antibody responses for rapid diagnostic tests. The inclusion of additional time points, greater sample size and asymptomatic cases are necessary to test the cut-off sensitivity scores; and to confirm the diagnostic value of our LIPS assays and the relevancy of ORF3b and ORF8 antibodies. Late-convalescent sera are needed to assess antibody waning. We also need to investigate antibody profiles with disease severity.

In conclusion, we found that COVID-19 patients not only produce antibodies to the Spike protein, but also to other structural and non-structural proteins. The nucleoprotein, and th en ORF8, and ORF3 show an immunodominant and specific humoral response compared with other SARS-CoV-2 antigenic targets tested. The combined use of N+ORF3b+ORF8 provides a sensitive and specific method for the detection of all COVID-19 patients in our cohort even at early time-points, whilst the Spike protein does not. Our results provide insights into the overall spectrum of antibody responses associated with COVID-19. We still need to investigate whether antigens other than the virus spike confer protection. Such information will help prioritize antigen targets for vaccine development, monoclonal antibody reagents and detecting early responses to infection.

## Methods

### Patients and samples collection

Our study enrolled a total of 26 patients with RT-PCR confirmed COVID-19 infection (Table 1). Fifteen patients were enrolled in Hong Kong (China, SAR) and all of them provided informed consent. The study was approved by the institutional review board of the Hong Kong West Cluster of the Hospital Authority of Hong Kong (approval number: UW20–169). Plasma samples were collected from heparinized blood, and heat-inactivated at 56°C, 30 mins. Gender- and age-matched plasma samples from healthy subjects collected before the COVID-19 pandemic were used as negative controls.

### SARS-CoV-2 gene cloning

Based on previous studies describing the structure of the SARS-CoV-2 genome (1, 8), an extensive panel of 12 proteins (S, E, M, N, NSP1, ORF3a, 3b, 6, 7a, 7b, 8, 10) was chosen for antibody testing by LIPS. Primers for the amplification of SARS-CoV-2 proteins were designed (See protein ID in Table 1, and primers sequences in Supplementary table 2). A RT-PCR was performed using extracted SARS-CoV-2 vRNA to amplify target genes corresponding to Structural and Non-structural proteins of the virus (Table 1, (6)) using Platinum SuperScriptIII One Step RT-PCR system. The bands were then extracted using Qiagen gel extraction kit (Qiagen, Germany) and digested with BamHI and NotI or KpnI-HF and XhoI (New England Biolabs, USA). Extracted products were ligated using T4 DNA ligase (New England Biolabs) into the pREN2 plasmid (from Peter Burbelo NDICR, NIH). Plasmids were transformed using DH10B competent cells and purified using PureYield Plasmid midi-prep system (Promega).

### SARS-CoV-2 (Ruc)-antigen expression

Constructs with pREN2-Renilla luciferase plasmid containing the SARS-CoV-2 antigen of interest were transfected into Cos1 cells using Fugene 6 (Promega) as per manufacturer’s instructions. Cells were harvested 48 hours later, lysed and sonicated, and (Ruc)-antigen yields were measured using a Luminometer plate reader (PerkinElmer) according to the protocol of Burbelo *et al*. (20).

### Measurement of antibody responses using Luciferase Immunoprecipitation System (LIPS)

The LIPS assays were performed following the protocol of Burbelo *et al*, with the following modifications (20). Briefly, (Ruc)-antigen (1e7 per well) and plasma (heat inactivated and diluted 1:100) were incubated for 2 hours with shaking at 800rpm. Ultralink protein A/G beads were added to the (Ruc)-antigen and serum mixture in a 96-deep-well polypropylene microtiter plate and incubated for 2 hours with shaking at 800rpm. The entire volume was then transferred into HTS plates and washed as previously described. The plate was read using QUANTI-Luc Gold substrate (Invivogen, rep-qlcg5) as per manufacturer’s instructions and a MicroBeta JET luminometer (PerkinElmer). Experimental controls include blank wells with antigens and negative control serum from age matched non-infected patient plasma collected prior to the COVID-19 pandemic.

### Enzyme-linked immunosorbent assay

ELISA assays were performed with the available SARS-CoV-2 proteins Spike (S1+S2) and Nucleoprotein (N) proteins. Briefly, recombinant S and N proteins (Sinobiological) were coated on 96-well flat-bottom immunosorbent plates (Nunc Immuno MaxiSorp, Roskilde, Denmark) at a concentration of 100 ng/ml, in 100/µl coating buffer (PBS with 53% Na_2_CO_3_ and 42% NaHCO_3_, pH 9.6) at 4°C overnight. An additional plate coated with a nonspecific protein (blocking buffer, PBS with 5% FBS) was used to measure the background binding of each sample. Following FBS blocking and thorough washing, diluted plasma samples (1:100) were bound for 2 hours, further washed and then detected by an anti-human Ig secondary antibody labelled with HRP specific for IgG (Invitrogen, Carlsbad, CA, USA).

### Microneutralization assay

Plasma samples were diluted in serial two-fold dilutions commencing with a dilution of 1:10, and mixed with equal volumes of SARS-CoV-2 at a dose of 200 tissue culture infective doses 50% (TCID_50_) determined by Vero E6 cells respectively. After 1 hour of incubation at 37°C, 35µl of the virus-serum mixture was added in quadruplicate to Vero or Vero E6 cell monolayers in 96-well microtiter plates. After 1 hour of adsorption, the virus-serum mixture was removed and replaced with 150μl of virus growth medium in each well. The plates were incubated for 3 days at 37°C in 5% CO_2_ in a humidified incubator. Cytopathic effect was observed at day 3 post-inoculation. The highest plasma dilution protecting 50% of the replicate wells was denoted as the neutralizing antibody titer. A virus back-titration of the input virus was included in each batch of tests.

### Multiple alignments of Coronaviruses

Multiple amino acid alignments of structural proteins from HKU1 (AY597011.2), HCoV-229E (AF304460.1), HCoV-OC43 (AY391777.1) and HCoV-NL63 (AY567487.2) were compared versus SARS-CoV-2 (MN908947) using CLUSTAL 2.1.

### Statistics

GraphPad Prism 6 software (San Diego, CA) was used for statistical analysis. Antibody levels are presented as the geometric mean +/− standard deviation (stdev). For the calculation of sensitivity and specificity, cut-off limits for each antigen were derived from the mean value plus three standard deviations of the controls. Non-parametric Mann–Whitney U tests were used to compare the antibody levels between COVID-19 and negative groups, using the GraphPad 8 Prism software.

## Data Availability

The data of this study is available upon request.

## Acknowledgements

This study was partly supported by the Research Grants Scheme (GRF 17113718). We thank Linfa Wang (Duke NUS, Singapore) for technical advice for establishing the LIPS assay.

